# Excitation-Inhibition Balance in Schizophrenia Spectrum Disorders: EEG Criticality Reflects Frontal Metabolites and a Potential Compensatory Mechanism

**DOI:** 10.64898/2026.06.12.26354984

**Authors:** Genc Hasanaj, Marcel S. Kallweit, Berkhan Karslı, Verena Meisinger, Emanuel Boudriot, Lukas Roell, Julian Melcher, Gizem Vural, Enrico Schulz, Nicole Klimas, Susanne Schmölz, Matin Mortazavi, Maxim Korman, Alexandra S. Hisch, Deniz Yılmaz, Johanna Spaeth, Antonia Šušnjar, Lenka Krčmář, Joanna Moussiopoulou, Vladislav Yakimov, CDP Working Group, Michael J. Ziller, Oliver Pogarell, Andrea Schmitt, Alkomiet Hasan, Peter Falkai, Florian J. Raabe, Elias Wagner, Daniel Keeser

## Abstract

**Background:** The excitation-inhibition (E-I) balance is essential for normal brain functioning, while deviations from this balance have been implicated in several psychiatric disorders. However, the extent to which electroencephalography (EEG) and proton magnetic resonance spectroscopy (¹H-MRS) E-I markers are altered in schizophrenia spectrum disorders (SSD), how they converge across modalities, and how they relate to cognitive performance and clinical symptoms remain insufficiently characterized.

**Methods:** We recruited 111 healthy controls (HC) and 113 individuals with SSD. All participants underwent resting-state EEG and ¹H-MRS. Metabolites were measured either in the anterior cingulate cortex (ACC; N_SSD_ = 63, N_HC_ = 58) or in the left dorsolateral prefrontal cortex (lDLPFC; N_SSD_ = 50, N_HC_ = 53), from which gamma-aminobutyric acid (GABA), glutamate + glutamine (Glx), and the Glx/GABA ratio were extracted. Extracted EEG E-I markers included oscillatory activity, aperiodic activity, functional E-I, microstates, multiscale entropy, and neuronal avalanche criticality.

**Results:** MRS results showed no group differences in GABA, Glx, or the Glx/GABA ratio. In contrast, most EEG-derived E-I markers indicated increased cortical inhibition in SSD, including steeper aperiodic exponents, prolonged microstate durations, and greater prevalence of subcritical states. However, functional E-I showed a divergent pattern, suggesting balanced dynamics in SSD and relatively inhibition-weighted dynamics in HC. Across groups, higher ACC and lDLPFC GABA predicted a lower kappa index, whereas a higher lDLPFC Glx/GABA ratio was associated with a higher kappa index. In SSD, reduced avalanche criticality was associated with better cognition and less severe symptoms.

**Conclusion:** Several EEG-derived E-I proxies, but not MRS measures, indicate an increased cortical inhibition in SSD. Criticality indices best capture frontal neurochemical metabolites and improvements in clinical symptoms, potentially reflecting inhibitory compensation mechanisms in SSD.

## 1. Introduction

The balance between excitatory and inhibitory (E-I) neural activity represents a fundamental feature of the brain that supports efficient information processing, network stability, and adaptive behavior^1,2^. The coordinated interplay between excitatory glutamatergic pyramidal neurons and inhibitory gamma-aminobutyric acid (GABA) interneurons is suggested to maintain this balance by regulating the strength, timing, and synchrony of cortical activity^3,4^. Failure to maintain this balance, whether through excessive inhibition or excitation, has been implicated in the pathophysiology of several neuropsychiatric disorders^5^.

Abundant postmortem, molecular, and neuroimaging evidence suggests a disruption of the E-I mechanism in schizophrenia spectrum disorders (SSD)^6^. Specifically, SSD patients show abnormal GABAergic signaling involving parvalbumin interneurons and N-methyl-D-aspartate receptor function^6^. Alterations in E-I balance have been shown to vary across illness stages and symptom severity, providing a potential stage-sensitive marker of pathophysiological progression in SSD^7,8^. An in-depth understanding of these markers and their role in SSD symptomatology might pave the way for future interventions^9,10^.

Several neuroimaging markers have been proposed to reflect the neural E-I dynamics. Among those, magnetic resonance spectroscopy (MRS) provides a non-invasive approach to quantify metabolite concentrations related to inhibitory and excitatory neurotransmission, namely GABA, glutamate, and glutamine^11^. On the other hand, several electrophysiological markers of electroencephalography (EEG) are proposed to index underlying E-I dynamics^12^. Identifying reliable EEG-based E-I markers could provide valuable clinical utility considering their comparatively inexpensive, scalable, and suitable usage for repeated measures, positioning them as promising markers for stratification and longitudinal monitoring of SSD^13,14^.

Although the EEG and MRS markers are frequently proposed and interpreted within the E-I framework, it remains unclear whether MRS- and EEG-derived E-I markers reflect an interconnected biological mechanism, capture complementary levels of dysregulation, or differ in their relevance to cognition and clinical symptoms in SSD. To address this, the present study adopts an exploratory EEG-MRS approach to investigate these proposed E-I markers in both healthy controls (HC) and SSD. Specifically, we examine whether frontal metabolites from the anterior cingulate cortex (ACC) and left dorsolateral prefrontal cortex (lDLPFC), together with EEG E-I markers, (i) differ between groups, (ii) converge across modalities, and (iii) relate to cognitive performance and clinical symptom severity in SSD. By integrating MRS and EEG, we aim to provide an exploratory characterization of E-I dysregulation in SSD and to investigate whether neurochemical and electrophysiological measures converge in indexing altered E–I balance.

## 2. Materials and Methods

### 2.1 Study Sample, Design, and Assessments

All 224 participants (113 SSD patients and 111 HC) in this study were recruited through the cross-sectional Clinical Deep Phenotyping (CDP) study^15^. The study was approved by the Ethics Committee of the Faculty of Medicine, LMU Munich (project numbers: 20-0528 and 22-0035). Participants were aged between 18 and 65 years. The patients were recruited from the inpatient and outpatient populations of the Department of Psychiatry and Psychotherapy at LMU University Hospital, whereas healthy controls were recruited through advertisements (e.g., flyers and online postings). All participants provided their informed consent. Patients were eligible if they had a diagnosis of schizophrenia (SZ), schizoaffective disorder (SZA), brief psychotic disorder (BrPsyD), or delusional disorder (DD) according to the Mini-International Neuropsychiatric Interview (M.I.N.I.)^16^. The majority of patients in this study received treatment with second-generation antipsychotics. Exclusion criteria included any primary psychiatric disorder apart from those specified; relevant past or present central nervous system diseases (such as multiple sclerosis, epilepsy, history of encephalitis, stroke, or cerebral surgery); pregnancy; significant language barriers; and substance use (except for alcohol, nicotine, and caffeine) more than once over the past 12 months. Trained staff assessed symptom severity using the Positive and Negative Syndrome Scale^17^ (PANSS) and psychosocial functioning using the Global Assessment of Functioning (GAF) scale. Cognitive performance was evaluated with the Brief Assessment of Cognition in Schizophrenia (BACS)^18^. Clinical remission was determined using the modified Andreasen criteria without the time criterion^19^. All participants completed resting-state EEG and magnetic resonance spectroscopy (MRS) in separate sessions (median interval: 0 days, range: 0-83 days). See Table 2 for combined cohort characteristics, as well as Tables S1 and S2 for cohort characteristics separated based on MRS region acquisition.

### 2.2 ^1^H-MR Spectroscopy Acquisition, Processing, and Postprocessing Quality Control

All participants underwent cranial magnetic resonance imaging (MRI) scanning at the Neuroimaging Core Unit Munich (NICUM) on a Siemens 3-Tesla Magnetom Prisma scanner (Siemens Healthcare, Erlangen, Germany) with a 32-channel head coil. For the MRI examination, participants were positioned at the scanner, provided with hearing protection, and instructed to remain still for the duration of the acquisition. T1-weighted images were acquired using a magnetization-prepared rapid acquisition gradient echo (MPRAGE) sequence (slice thickness: 0.8 mm, repetition time: 2500 ms, echo time: 2.22 ms, FOV: 256 mm^2^, flip angle: 8°).

Each participant underwent a single-voxel proton magnetic resonance spectroscopy (^1^H-MRS) measurement. Trained MRI operators used T1w images for anatomical guidance and followed defined anatomical landmarks to manually place either a 20 × 20 × 20 mm^3^ sized volume of interest (VOI) in the anterior cingulate cortex (ACC; N_SSD_ = 63, N_HC_ = 58), or a 30 × 30 × 15 mm^3^ sized VOI in the left dorsolateral prefrontal cortex (lDLPFC; N_SSD_ = 50, N_HC_ = 53) (Figure S1 in Supplement 1). Metabolite acquisition from these two regions was already preselected in the parent (CDP) study^15^. Before spectroscopy acquisition, a fastestmap sequence was performed to optimize B0 shimming and the magnetic field homogeneity. All ^1^H-MRS measurements were conducted by a Mescher-Garwood semi-localized by adiabatic selective refocusing (MEGA-sLASER) sequence (repetition time: 3000 ms, echo time: 68 ms, editing pulse ON: 1.90 ppm, editing pulse OFF: 7.50 ppm, editing pulse bandwidth: 100 Hz, flip angle: 180°)^20^. This sequence enabled improved spectral separation of GABA from creatine and optimized the detection of low-concentration GABA^21–23^. For each MRS measurement, a water-suppressed scan with 128 averages was performed using variable power radiofrequency pulses with optimized relaxation delays (VAPOR)^24^. This was followed by a non-water-suppressed scan with 8 averages, which served as a reference for correcting the baseline and optimizing background noise estimation.

All spectroscopic data were processed using Osprey^25^ (v.2.5.0) and LCModel^26^ (Linear Combination Model v.6.3-1R). Preprocessing was conducted in Osprey and included motion and frequency drift corrections by using eddy current correction, rejection of motion-affected averages, and robust spectral alignment. Voxel placement consistency was visually inspected using images generated by Osprey’s CoRegistration feature. All preprocessed spectra were analyzed in LCModel employing a basis set specifically tailored for the exact MEGA-sLASER sequence used in this study at the NICUM site scanner. The basis set was created via numerical simulations in MATLAB, using the FID-A toolkit^27^. GABA was derived from the DIFF spectrum, whereas Glx was derived from the OFF spectrum. Finally, metabolite concentrations (millimolar, mM) were corrected for the cerebrospinal fluid (CSF) fraction within the VOI using individual T1w MRI segmentation obtained by Osprey, which utilizes SPM12 (http://www.fil.ion.ucl.ac.uk/spm/software/spm12/) for tissue segmentation. Metabolite concentrations were evaluated for data quality using multiple criteria. Values with LCModel estimated standard deviations based on Cramér-Rao lower bound (CRLB) greater than 30%, a signal-to-noise ratio less than 3, or a full-width half-maximum (FWHM) greater than 0.1 were replaced with missing values (NA)^28^. Furthermore, outlier metabolite concentrations extending more than three interquartile ranges beyond the third quartile or beneath the first quartile were identified and set to NA^29^. Additionally, spectral fits were assessed by visual inspection; however, no further exclusions were necessary (see Table S3 in Supplement 2 for mean MRS quality metrics). The final selected metabolites in this study included GABA, Glx (Glutamine + Glutamate), and Glx/GABA ratio.

### 2.3 Resting-State EEG Recording and Preprocessing

For the recordings, all participants were seated in a quiet, dimly lit study cabin and underwent EEG using a standardized 32-channel device (BrainAmp amplifier, Brain Products, Gilching, Germany) at the EEG laboratory of the Department of Psychiatry at LMU Munich. An Electro-Cap (Electro-Cap International, Inc, Eaton, OH, USA) was used following the International 10-20 system nomenclature (including two mastoid electrodes). Trained operators placed the electrodes and instructed the subjects to remain as relaxed and calm as possible to minimize ocular and muscular artifacts. The recording of 5 minutes of eyes-closed condition (with visual blockage reaction of 3 seconds after 2 minutes to prevent excessive alpha activity) was performed with a sampling frequency of 1000 Hz and Cz as the reference electrode, while the skin impedance was kept below 5 kΩ.

EEG data were preprocessed in EEGLAB^30^ (v2025.0), utilizing an automated in-home preprocessing pipeline as previously used by Adams et al.^31^. Initially, the data were re-referenced to the mastoid electrodes (A1 and A2) and then resampled to 256 Hz. A bandpass filter of 1–70 Hz was applied, and data were segmented into 6-second epochs. Each of these epochs was further preprocessed by removing linear trends and very low or high-frequency bands, rejecting channels based on atypical power spectrum values, joint probability, and kurtosis. Artifacts were removed using the Multiple Artifact Rejection Algorithm^32^ (MARA). Afterwards, the data underwent a second round of similar channel and epoch rejection, with only slight modifications. Rejected channels were interpolated at the end of preprocessing. Participants who did not meet the criteria for a sufficient number of epochs or non-rejected channels were excluded from further analyses (see Supplemental Methods in Supplement S1 for a detailed preprocessing description).

### 2.4 EEG-Derived Markers of E-I Balance

The post-processing of EEG data was conducted in Python and Matlab (The MathWorks, Inc). All EEG-EI proxies were selected from the comprehensive review of Ahmad et al.^12^, which identified several potential EEG markers that can reflect shifts in E-I activity (Table 1).

**Table 1.** Overview of selected EEG-derived excitation-inhibition (E-I) proxies and their proposed interpretation.

| EEG E-I proxy | Measure extracted | Method / frequency range | E-I interpretation |
| --- | --- | --- | --- |
| Gamma power and center frequency | Aperiodic-adjusted and canonical absolute gamma power and aperiodic-adjusted gamma center frequency | Canonical gamma power was extracted from PSD estimated with Welch's method, whereas aperiodic-adjusted gamma power and center frequency were extracted using *specparam*<sup>33</sup>; extraction range: 30–70 Hz. | Interpretation is directional and relative across groups/conditions. Gamma power is proposed to increase with a larger drive of inhibitory interneurons<sup>3,34</sup>. Similarly, gamma center frequency is suggested to increase as the network E-I balance shifts toward greater relative inhibition<sup>35</sup>. |
| Beta power | Aperiodic-adjusted and canonical absolute beta power | Aperiodic-adjusted and canonical absolute beta power were extracted using the same method as Gamma power within 12–30 Hz. | Interpretation is directional and relative across groups/conditions. Beta power is suggested to increase due to a larger drive of inhibitory interneurons<sup>36</sup>. |
| Aperiodic exponent | Exponents from 3–40 Hz, 30–48 Hz, and 1–70 Hz fitting ranges | Fixed *specparam* model fitted across three frequency ranges. | Interpretation is directional and relative across groups/conditions. Lower exponent values are suggested to index inhibition-dominated dynamics, whereas higher values suggest a relative shift towards excitation-dominated dynamics<sup>37,38</sup>. |
| Functional E-I (fEI) ratio | fEI value | DFA-based fEI computed in the extended alpha range, 6–13 Hz; only values with DFA between 0.55 and 1 retained. | fEI $\approx 1$ indicates balanced E-I; fEI $< 1$ indicates inhibition-dominated/subcritical dynamics; fEI $> 1$ indicates excitation-dominated/supercritical dynamics<sup>39</sup>. |
| Microstate dynamics | Mean durations and occurrences for microstate classes A–G | Seven-class microstate solution; EEG filtered 2–20 Hz; MICROSTATELAB/EEGLAB<sup>40</sup>. | Increased microstate durations may reflect more rigid, less flexible, and potentially inhibition-weighted network dynamics<sup>41,42</sup>. |
| Multiscale sample entropy (MSE) | MSE across full-range, low-frequency, alpha, beta, and gamma scale ranges | NeuroKit2<sup>43</sup> multiscale entropy; embedding dimension $m = 2$, tolerance $r = 0.2$; scale ranges approximating full-range, low-frequency, alpha, beta, and gamma bands. | Peak entropy has been proposed to reflect maximal neural state repertoire in balanced E-I networks operating near criticality<sup>44,45</sup>. |
| Neuronal avalanches | Avalanche exponent, branching ratio, and kappa ($\kappa$) index | Avalanche bin size optimized to approximate $\alpha = -3/2$ in healthy controls. Branching ratio and avalanche exponent were calculated to assess network excitability, and kappa index was computed to evaluate the goodness of fit to the theoretical $\alpha = -3/2$ distribution based on Shew et al.<sup>46</sup> | $\kappa \approx 1$ indicates criticality; $\kappa < 1$ indicates inhibition-dominated/subcritical dynamics; $\kappa > 1$ indicates excitation-dominated/supercritical dynamics<sup>46,47</sup>; higher/lower branching ratios and avalanche exponents indicate a shift toward excitation-/inhibition-dominated supercritical/subcritical dynamics, respectively<sup>47,48</sup>. |

**Table 2.**
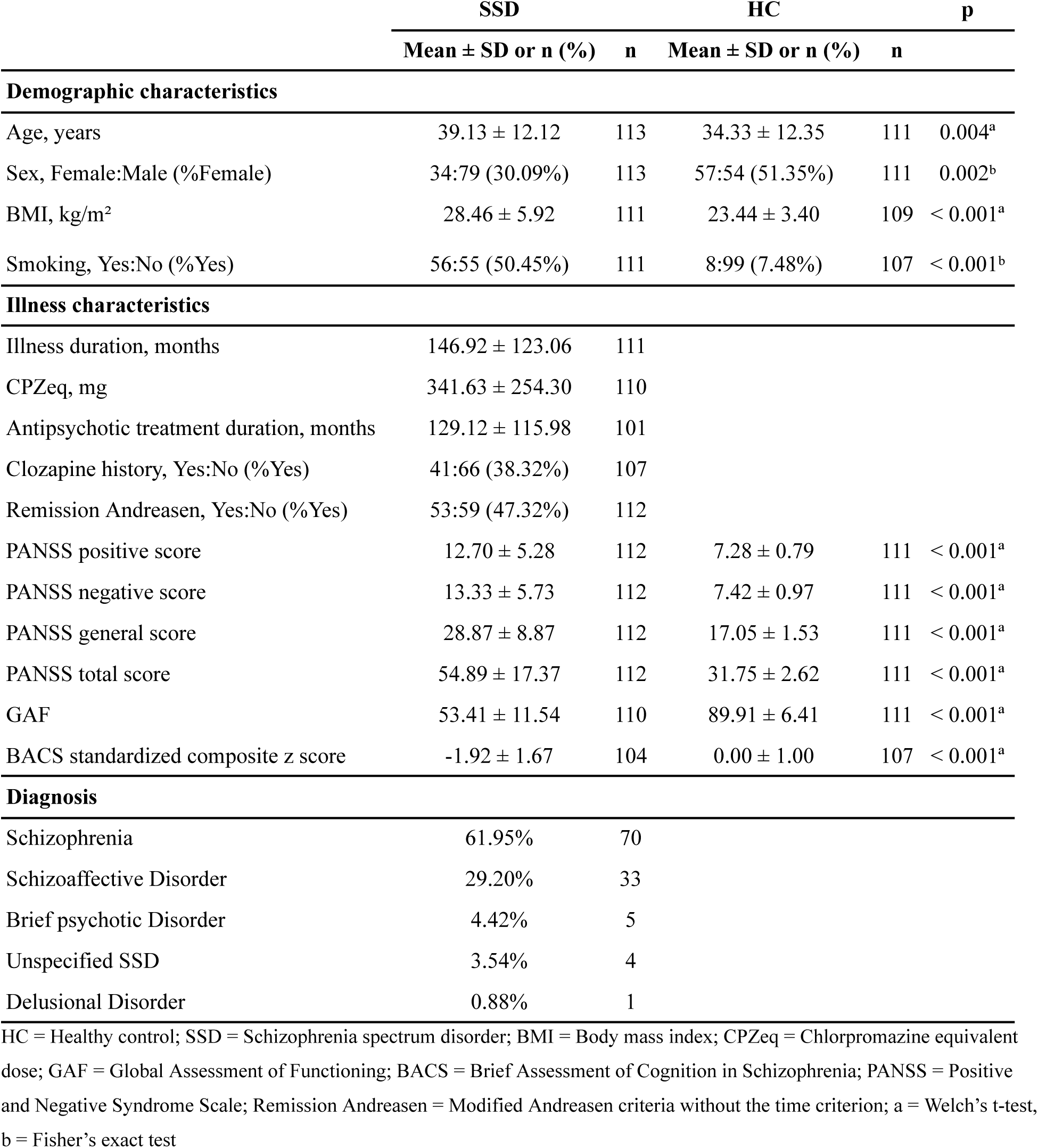
Sample characteristics.

The EEG-based E-I proxies were evaluated at two spatial scales. We defined global-level metrics as those derived by either averaging activity across all electrodes or computing the measure using the collective activity of the entire array of electrodes (i.e., microstates and neuronal avalanches). Global-level measures included gamma power (canonical and aperiodic-adjusted), gamma center frequency, beta power (canonical and aperiodic-adjusted), aperiodic exponents (3–40 Hz, 30–48 Hz, and 1–70 Hz), fEI, microstates (7-class solution map for microstate duration and occurrence), neuronal avalanches (avalanche exponent, branching ratio, and kappa index), and multiscale sample entropy (low-frequency, alpha, beta, gamma, and full ranges). Considering the regional relevance of certain markers^12^, we also performed electrode clustering by averaging sensor-level values across predefined scalp regions (frontal: FP1, FP2, F3, F4, Fz; central: C3, C4, Cz, FC1, FC2; left temporal: F7, FC5, T3, CP5, T5, FT9; right temporal: F8, FC6, T4, CP6, T6, FT10; parietal: P3, P4, Pz, P9, P10; occipital: O1 and O2). Regional cluster measures included all proposed resting-state EEG E-I proxies except microstate dynamics and neuronal avalanches, which were assessed exclusively at the global level.

### 2.5 Statistical Analyses

All statistical analyses were conducted in R^49^. The analyses were separated into two main steps: testing for diagnostic group differences and testing for cross-modal relationships. To assess group differences in metabolite concentrations and global EEG-derived E-I proxies we used analysis of covariance (ANCOVA), with the diagnostic group entered as the main factor. For regional EEG measures, we used linear mixed-effects models (LMMs; *lme4*^50^ and *lmerTest*^51^). In these models, the regional EEG metric was the dependent variable. Diagnostic group, electrode cluster, and their interaction were entered as fixed effects. Participant was included as a random intercept to account for repeated measurements across electrode clusters. Next, to test whether MRS-derived neurochemical markers predicted EEG-based E-I proxies, we used linear regression for the global EEG metrics. The global EEG metric was specified as the dependent variable, with the MRS metabolite, diagnostic group, and their interaction entered as predictors. For regional EEG metrics, we used LMMs. The regional EEG metric served as the dependent variable, while the MRS measure, diagnostic group, electrode cluster, and all their interaction terms were specified as fixed effects, alongside a random intercept for participants. Age and sex were used as covariates for all the models.

Type III tests were used throughout. For linear models, omnibus effects were obtained from Type III sums of squares, whereas for mixed models, omnibus effects were evaluated using Type III Wald F-tests with the Kenward–Roger approximation for denominator degrees of freedom^52^. Significant omnibus effects were followed by post-hoc analyses based on estimated marginal means, implemented using the *emmeans* package. For interaction terms involving continuous predictors, follow-up analyses were performed using estimated marginal trends (simple slopes), together with pairwise comparisons of slopes where appropriate. To control for multiple testings, p-values were adjusted using the Benjamini–Hochberg false discovery rate (FDR) method^53^ (q value), while grouping all the corrections per domain (oscillations, aperiodic activity, microstates, avalanche criticality, multiscale sample entropy, and functional E-I). For non-mixed models, effect sizes were summarized using partial eta squared (*η*²7) with confidence intervals, as implemented in the effectsize^54^ package. The significant level was set at a q value < 0.05.

Partial correlation analyses were conducted in SSD to examine the relationships of all EEG and MRS E-I proxies with cognitive (BACS composite score) and clinical symptom severity (PANSS positive, PANSS negative, PANSS total), controlling for age, sex, and chlorpromazine equivalent (CPZeq) dose effects.

## 3. Results

### 3.1 ^1^H-MRS metabolite comparison between HC and SSD

No significant differences between SSD and HC were found for GABA, Glx, or Glx/GABA ratio in either the ACC or the lDLPFC, all q > 0.05 (Figure S4 in Supplement 1 and Table S4 in Supplement 2).

### 3.2 Case-control differences across different proposed EEG E-I markers

At the global level, SSD participants showed steeper aperiodic exponents than HC participants across both the 1–70 Hz, F(1, 218) = 13.275, p < .001, q < .001, η²7 = .057, and the 3–40 Hz fitting range, F(1, 218) = 12.479, p < .001, q < .001, η²_p_ = .054. The same direction of effect was observed across all electrode clusters, with all cluster-level comparisons surviving correction for multiple comparisons, all q < .05. SSD participants showed higher global functional E-I (fEI), F(1, 217) = 8.404, p = .004, q = .004, η²_p_ = .037, with significant effects in central, left and right temporal, parietal, and occipital clusters, all q < .05. Although the 30–48 Hz aperiodic exponent did not differ significantly at the global level, F(1, 218) = 3.861, p = .051, q = .051, η²_p_ = .017, electrode-cluster analyses revealed a significant group × cluster interaction, F(5, 1100) = 2.852, p = .015, q = .015. Post hoc analyses indicated a localized increase in the frontal cluster in SSD compared with HC, B = 0.348, t = 2.76, p = .006, q = .036 (Figure 2b).

SSD participants also showed significantly prolonged microstate durations compared with HC, particularly for microstate classes A, F(1, 210) = 9.178, p = .003, q = .019, η²_p_ = .042; B, F(1, 210) = 7.734, p = .006, q = .027, η²_p_ = .036; C, F(1, 210) = 6.197, p = .013, q = .038, η²_p_ = .029; and E, F(1, 210) = 5.420, p = .021, q = .048, η²_p_ = .025. In addition, SSD participants showed increased beta-range multiscale entropy, F(1, 218) = 8.635, p = .003, q = .018, η²_p_ = .038, with significant effects in left and right temporal, parietal, and occipital electrode clusters, all q < .05.

By contrast, SSD participants showed lower avalanche exponent, F(1, 218) = 10.894, p = .001, q = .002, η²_p_ = .048; branching ratio, F(1, 218) = 24.772, p < .001, q < .001, η²_p_ = .102; and κ index, F(1, 218) = 7.708, p = .006, q = .006, η²_p_ = .034. SSD participants also showed reduced occurrence of microstate classes D, F(1, 210) = 14.886, p < .001, q = .002, η²_p_ = .066, and G, F(1, 210) = 6.722, p = .012, q = .035, η²_p_ = .030 (Figure 2a; Tables S5–S7 in Supplement 2).

Additional significant group × electrode-cluster interactions were observed. For canonical gamma power, the group × electrode cluster interaction was significant, F(5, 1100) = 6.743, p < .001, q < .001. Although the mean of gamma power was lower in SSD across all electrode clusters, the post hoc comparison survived correction only in the occipital cluster, B = −0.334, t = −3.32, p = .001, q = .008. For low-frequency multiscale entropy, the group × cluster interaction was also significant, F(5, 1100) = 4.128, p = .001, q = .001, with localized decreases in SSD in the frontal cluster, B = −0.070, t = −2.864, p = .005, q = .023, and central cluster, B = −0.066, t = −2.685, p = .008, q = .023. Although further group × cluster interactions were observed for beta power, both canonical and aperiodic-adjusted, alpha multiscale entropy, and full-range multiscale entropy, post hoc comparisons did not survive correction, all q > .05 (Figures 1c–d; Tables S6–S7 in Supplement 2).

**Figure 1.**
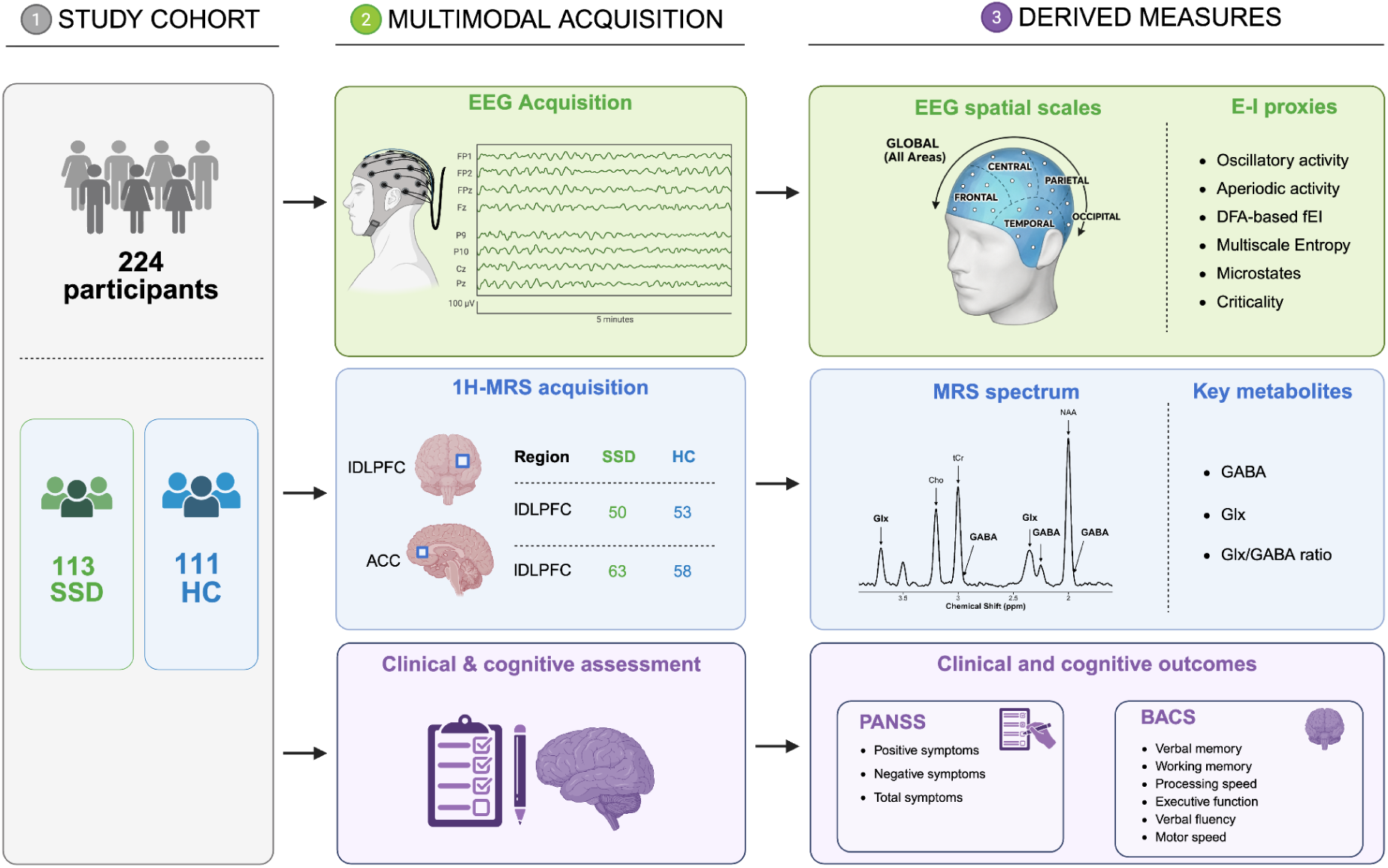
Study design and data acquisition overview.

**Figure 2:**
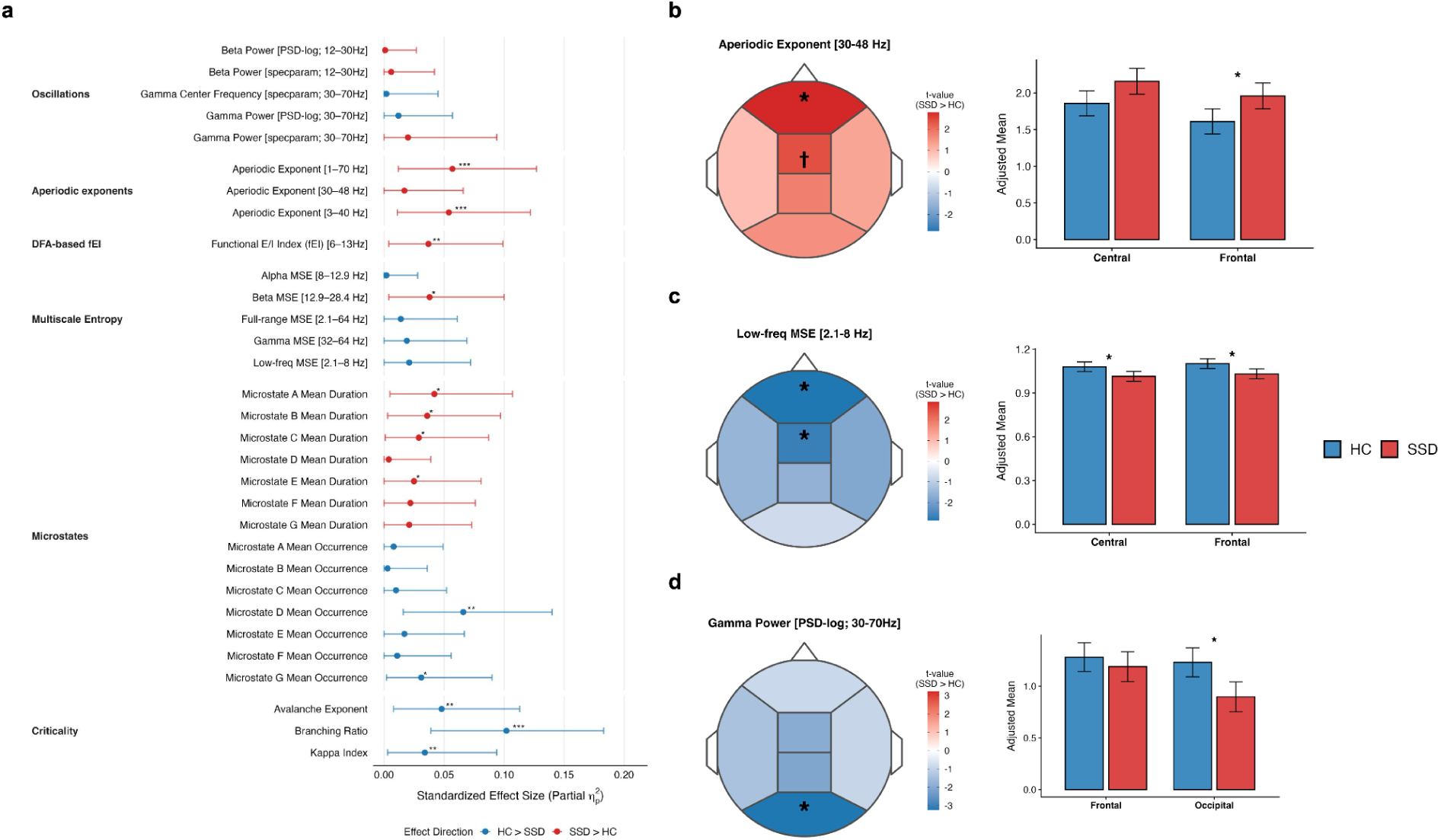
Global and regional case-control divergence in electrophysiological E-I proxies. **a** Forest plot illustrating the partial effect sizes (η²_p_) and their associated two-sided 95% confidence intervals (CIs) for case-control comparisons between healthy controls (HC) and individuals with schizophrenia spectrum disorders (SSD). Markers are stratified by their respective functional E-I domains. **b, c and d** Spatial and magnitude characterization of **(b)** the aperiodic exponent (30–48 Hz) and **(c)** low-frequency multiscale entropy (MSE; 2.1–8 Hz), and **(d)** canonical gamma power (30–70 Hz). The left panels depict scalp topographies (t-maps) of case-control contrast across predefined electrode clusters, highlighting the anterior-posterior distribution of the effect. The right panels show post-hoc analysis of Estimated Marginal Means (EMMs; adjusted for age and sex) for the two most dominant central and frontal electrode clusters, and the error bars represent 95% CIs. Significance thresholds are denoted by the following: † q < 0.10, * q < 0.05, ** q < 0.01, and *** q < 0.001.

### 3.3 Relationship between metabolite and electrophysiological E-I proxies

Multiple linear regression analyses examining associations between MRS metabolites and electrophysiological E-I proxies revealed significant associations between GABA and the neuronal avalanche κ index. Specifically, higher GABA levels in both the ACC, F(1, 50) = 10.382, p = .002, q = .013, and lDLPFC, F(1, 91) = 6.415, p = .013, q = .039, were associated with lower κ values. In addition, the lDLPFC Glx/GABA ratio was positively associated with the κ index, F(1, 91) = 7.338, p = .008, q = .049 (Figure 3a; Table S8 in Supplement 2). Cluster-level analyses further revealed significant metabolite × electrode-cluster interactions for Glx in relation to the 30–48 Hz aperiodic exponent, F(5, 455) = 3.328, p = .006, q = .035, and fEI, F(5, 462.04) = 3.260, p = .007, q = .013. Post hoc analyses indicated that higher Glx levels in both regions were associated with steeper fronto-central aperiodic slopes and higher fronto-central fEI relative to parieto-occipital clusters, all q < .05 (Tables S9–S10 in Supplement 2).

**Figure 3.**
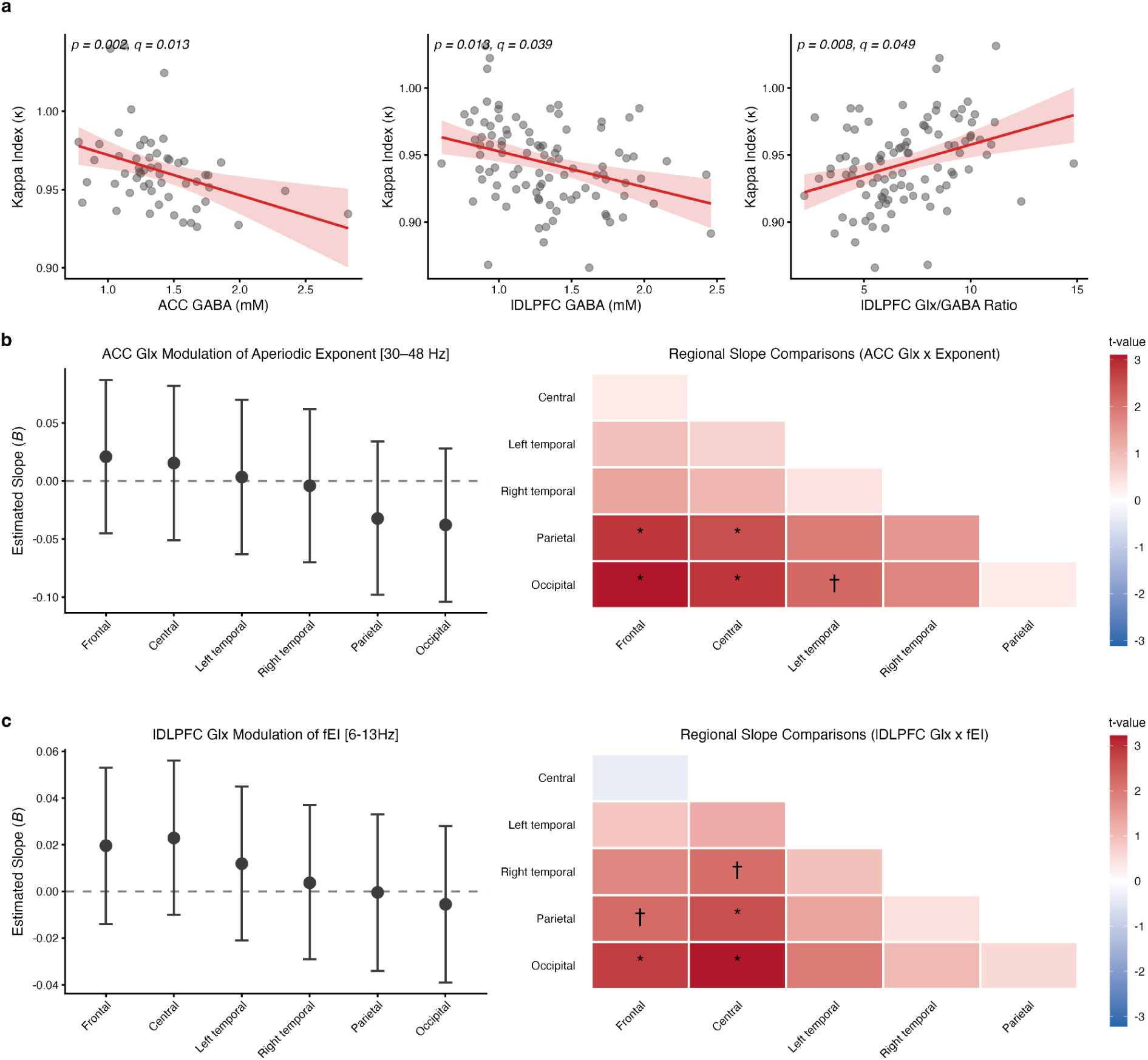
Neurochemical modulation of global neuronal criticality and regional electrophysiological E-I markers. a-b. Scatter plots depicting the significant continuous relationships between MRS metabolites and the global kappa index (κ), a proxy for neuronal avalanche criticality. **a** The left and middle panels show higher levels of anterior cingulate cortex (ACC) GABA and left dorsolateral prefrontal cortex (lDLPFC) GABA significantly predict reduced global κ (shifting towards the subcritical regime), respectively. The right panel, conversely, shows that a higher lDLPFC Glx/GABA ratio is associated with elevated κ. Solid red lines represent the linear model fit, with shaded regions indicating the 95% confidence intervals. **b-c** Visualization of the spatial gradients driving the significant Glx x Electrode Cluster interactions. Left panels depict point-range plots of the unstandardized slopes (B coefficients with 95% CI) estimating the effect of Glx on the respective EEG E-I proxy within each specific electrode cluster, ordered from anterior to posterior. The right panels show heatmaps of the pairwise regional slope comparisons. Colors represent the t-statistic of the contrast; red indicates that the region on the y-axis has a more positive Glx-EEG slope than the region on the x-axis. **c** ACC Glx modulation of the high-frequency aperiodic exponent (30–48 Hz) exhibits a distinct anterior-to-posterior gradient, with significantly steeper slopes in frontal/central clusters compared to parietal/occipital clusters. **c** A similar spatial gradient is observed for lDLPFC Glx modulation of the functional E-I Index (fEI; 6–13 Hz). All reported p-values are derived from the linear models, and significance markers denote False Discovery Rate (FDR)-corrected q-values: † q < 0.10, * q < 0.05.

### 3.4 Association between E-I proxies and cognitive and clinical symptoms

To further examine associations between clinical symptoms, cognitive performance, and EEG-and MRS-derived E-I proxies, we performed partial correlations between E-I markers and PANSS and BACS scores within the SSD group, controlling for age, sex, and CPZeq. At the global EEG level, only avalanche-criticality measures were significantly associated with clinical and cognitive outcomes. Specifically, higher branching ratio was associated with higher PANSS positive, *r*_partial_ = .306, 95% CI [0.12, 0.47], p = .002, q = .007; negative, *r*_partial_ = .226, 95% CI [0.04, 0.40], p = .020, q = .035; and total scores, *r*_partial_ = .289, 95% CI [0.10, 0.46], p = .003, q = .007. Higher κ index was associated with higher PANSS positive, *r*_partial_ = .277, 95% CI [0.09, 0.44], p = .004, q = .009, and total scores, *r*_partial_ = .217, 95% CI [0.03, 0.39], p = .026, q = .035. Higher avalanche exponent was associated with higher PANSS positive, *r*_partial_ = .222, 95% CI [0.03, 0.40], p = .023, q = .035; negative, *r*_partial_ = .289, 95% CI [0.11, 0.46], p = .002, q = .007; and total scores, *r*_partial_ = .306, 95% CI [0.12, 0.47], p = .002, q = .007, as well as lower BACS composite scores, *r*_partial_ = −.301, 95% CI [−0.47, −0.11], p = .003, q = .007 (Figure 4; Table S11 in Supplement 2). No significant association was found between regional EEG markers and clinical or cognitive performance (Figure S5 in Supplement 1).

**Figure 4.**
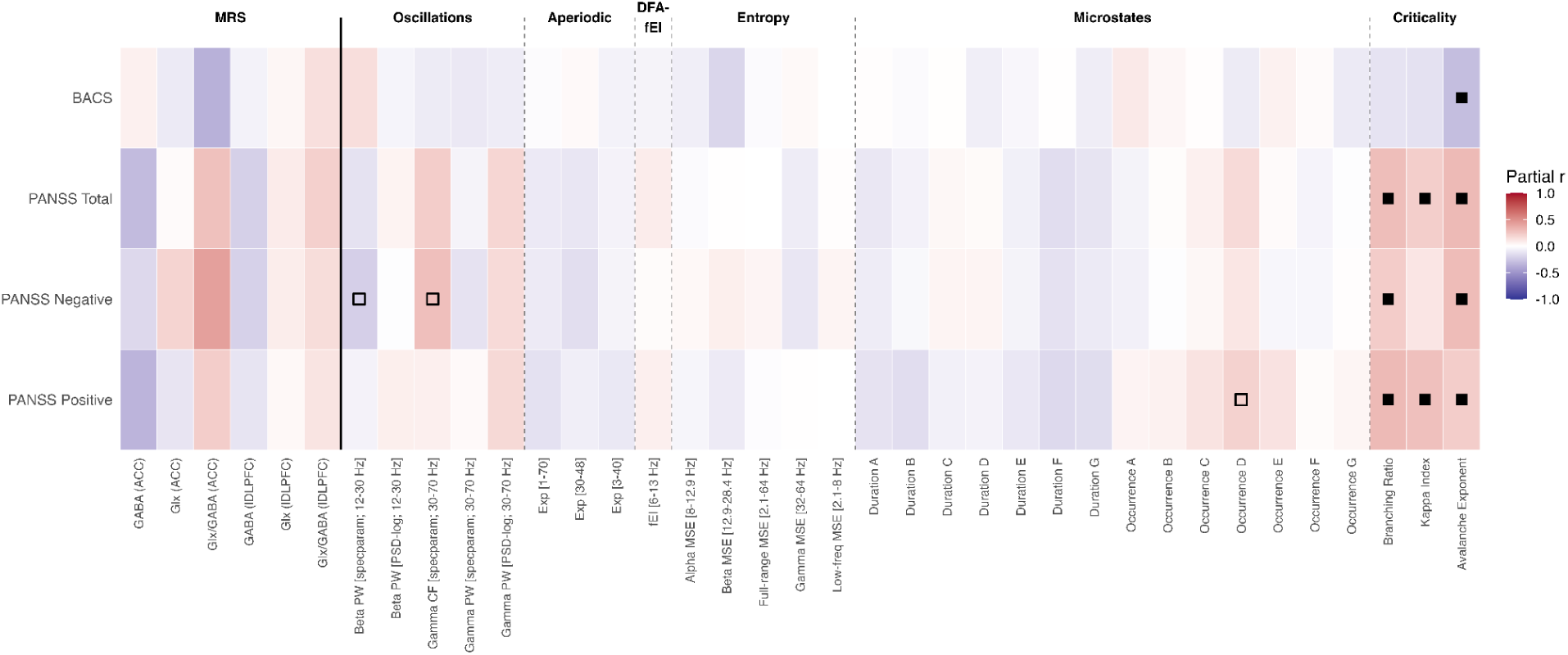
Partial correlations of region-specific MRS metabolites and electrophysiological E-I proxies with clinical and cognitive measures in schizophrenia spectrum disorders. Rows represent PANSS total, PANSS positive, PANSS negative, and BACS composite z-score. Columns include MRS metabolites from the ACC and left DLPFC, followed by EEG-based measures related to oscillatory activity, aperiodic activity, fEI, multiscale entropy, microstates, and neuronal avalanche dynamics. All partial correlations were adjusted for age, sex, and chlorpromazine-equivalent dose (CPZeq). Color indicates the strength and direction of the partial correlation, with positive values shown in green and negative values in purple. Open squares mark nominally significant associations (p < .05), and filled squares indicate associations that remained significant after FDR correction (q < .05).

## 4. Discussion

In this multimodal study, we assessed a broad array of putative E-I markers in HC and SSD, including MRS metabolites and EEG-derived E-I measures. Our findings can be summarized in four main points: (i) While traditional metabolite measures (GABA, Glx, and Glx/GABA derived from ACC and lDLPFC) did not differ between SSD and HC at the group level, (ii) several proposed EEG-based E-I indices revealed significant alterations in SSD, and (iii) the cross-modal relationship between MRS and EEG revealed associations between frontal GABA and Glx/GABA ratio with neuronal avalanche κ index. Furthermore, among patients, (iv) E-I dynamics reflecting increased inhibition via subcritical states were associated with less severe clinical symptoms and better cognitive performance.

### 4.1. No metabolite differences between HC and SSD in ACC and lDLPFC

The lack of metabolite differences in this study is broadly consistent with prior meta-analyses showing that MRS metabolite abnormalities in chronic and established SSD are generally small, heterogeneous, and often dependent on subgroup composition, illness stage, or analytic approach^55–59^. In addition, the absence of significant effects in the present study may reflect regional specificity and treatment-related influences. Specifically, previous works have suggested that metabolite abnormalities may be more evident in other medial frontal subregions, rather than in the rostral ACC examined in this study, and that antipsychotic exposure may attenuate earlier excitatory abnormalities^60–62^. Moreover, methodological variation across studies such as voxel placement, field strength, tissue correction, and macromolecule suppression likely contributes to the mixed and region-dependent pattern of findings in the literature^63,64^. Overall, our results support the view that MRS-derived metabolite alterations in predominantly chronic SSD are subtle rather than robust, and may vary according to region, clinical state, and treatment history.

### 4.2. Avalanche criticality reflects regional metabolite concentration and suggests inhibitory support in SSD

Brain criticality is thought to be tightly coupled with the brain’s E-I dynamics^12,65,66^. Neuronal avalanche dynamics provide a quantitative framework for characterizing how cascades of activity follow scale-free, power-law statistics, where the κ index was proposed as a sensitive electrophysiological marker of E-I balance by indexing a system’s proximity to criticality (κ = 1) versus subcritical (κ < 1) or supercritical (κ > 1) regimes^12,46,47^. Operating near criticality (slightly subcritical) is thought to support both efficient neural information transmission and capacity^46,67^, and is observed across different experimental settings^68–71^ and species^72,73^. The negative association between κ and GABA in both the lDLPFC and ACC, together with the positive association between κ and the Glx/GABA ratio in the lDLPFC, suggests that frontal neurochemical balance is coupled to large-scale electrophysiological dynamics. Specifically, higher GABA levels were associated with lower κ values, whereas a higher Glx/GABA ratio was associated with higher κ values. Our results align with previous studies showing that reduced inhibition can shift the system toward critical or supercritical regimes, as demonstrated in computational models^74,75^, in vitro pharmacological evidence^46^, and state-dependent studies across altered levels of consciousness^70^.

Under the E-I framework, the observed decrease in neural avalanche indices (κ index, branching ratio, and avalanche exponent) in our predominantly chronic, medicated, and relatively clinically stable sample of SSD may indicate a shift toward more subcritical and inhibition-weighted large-scale neural dynamics. E-I imbalance is considered a core physiopathology feature of SSD^9^. However, E-I imbalance may not represent a fixed trait of the illness, but rather a state-dependent process that varies across stages and clinical conditions^7^. Excessive excitation, for example, has been more frequently reported in earlier stages of the disorder, such as in clinical high-risk states and first-episode psychosis, and this increase is tightly linked with positive symptoms, while these effects appear attenuated or reversed at later stages^77–79^. The underlying mechanism of the global/background inhibitory activity in schizophrenia, as was shown with biophysical models, may be attributed to the reduced gain of pyramidal cells via increased self-inhibition^31^.

Considering the brain’s homeostasis tendency to restore imbalanced conditions of E-I^80^, the more subcritical and inhibition-weighted dynamics may reflect a later-stage compensatory response mechanism of the early-stage excitatory excess in SSD. Restored inhibitory dynamics may thus mirror the symptoms offset. In line with this, we observed a relationship of total symptoms in general, and positive symptoms in particular, with all avalanche measures. Respectively, a shift towards subcritical states in SSD across the three neuronal avalanche indices was associated with lower symptoms. Although the orientation towards subcritical states of these three indices pointed towards improved cognition, only the avalanche exponent showed a significant link. The current literature suggests a close link between critical states and cognition^67^. Specifically, slightly subcritical states in healthy controls are linked with a better focus of attention during a task^81^, meditation^82^, or fluid intelligence^83^. On the same line, in an overlapping sample from the current cohort, we found that higher left DLPFC GABA is associated with larger centroparietal P300 amplitudes in SSD, which in turn indexes a better cognitive performance^8^, further supporting the notion that inhibitory-dominated activity may reflect compensatory effects on cognition in later-stage and medicated SSD.

### 4.3. Additional electrophysiological proxies suggest altered E-I dynamics in SSD

In addition to brain avalanche criticality, two other electrophysiological markers, namely the aperiodic exponent and fEI, represent two other metrics closely associated with E-I dynamics. Computational and pharmacological evidence demonstrates that steeper aperiodic exponents are associated with stronger inhibitory neural activity^37,38^. Aligning with this hypothesis, and also with our previous neuronal avalanche findings, the robust evidence of steeper aperiodic exponent at different ranges (3–40 Hz; 1–70 Hz; 30–48 Hz) we found in SSD, may further support an inhibition-weighted brain activity in SSD. Unlike the aperiodic exponent, which does not provide a reference value for balanced E-I states, fEI has been proposed to index E-I dynamics relative to an interpretable reference point, with values around 1 indicating balance, values below 1 suggesting hypoexcitability, and values above 1 suggesting hyperexcitability.^39^. Under this framework, our current fEI findings deviate from previous E-I interpretation, with SSD individuals in almost balanced states (mean ≈ 1), but HC with a significant drop towards hypoexcitability (mean ≈ 0.9). Similar findings of hypoexcitability direction in HC versus a more balanced state in the clinical group (i.e., Alzheimer’s disease) have been documented in another study^84^, suggesting that this index may not straightforwardly capture balanced states in HC as originally proposed, and should therefore be interpreted with caution. While none of these markers were associated with metabolites, Glx derived from ACC association with aperiodic component (30–48 Hz), and of lDLPFC with fEI, showed a clear spatial gradient of increased slopes in the frontal regions and decreased slopes in posterior regions, suggesting a potential sensitivity of the markers for direct local effects of the regions in closer proximity to the MRS voxels. Although this interpretation remains speculative, other studies utilizing high-resolution source localization, as well as the 7T scanner, can better elucidate this relationship, as well as for other recommended EEG E-I proxies that may better represent local (im)balances.

Additional support for increased inhibition patterns in our study comes from another large-scale brain activity, namely microstate dynamics, that reflect quasi-stable functional configurations of the intrinsic electrophysiological activity^85,86^. Although not intrinsically indexing a state of excitation or inhibition, previous studies have shown that microstate durations are sensitive to pharmacological manipulations and non-invasive brain stimulation. Specifically, administration of lorazepam (a GABA agonist) has been found to reorganize and strengthen the dynamics of these quasi-stable microstates^41^. Similarly, inhibitory transcranial magnetic stimulation over the motor regions increased the duration of the microstates, inducing both acute and long-term effects^42^. Within this framework, the global microstate duration increases across all seven classes that we observed in SSD, although reaching statistical significance only for classes A, B, C, and E, might further reflect a shift towards inhibition-dominated brain dynamics in our sample. Importantly, microstate abnormalities may provide further evidence that E-I dynamics may shift during illness courses. A recent study in clinical high-risk individuals reported shorter microstate durations and increased occurrence, particularly in those who later converted to psychosis, suggesting reduced temporal stability at earlier stages of psychosis^87^. By contrast, our predominantly chronic and medicated SSD sample showed longer microstate durations, in line with meta-analytic evidence indicating increased duration in chronic SSD^88^. Together, these findings suggest that microstate dynamics in SSD may evolve, and potentially shift, across illness stages. Reduced temporal stability may reflect excessive excitation during early conversion stages^7^, which may later be compensated or overcompensated during the chronic course of the disorder. Although the absence of associations with MRS metabolites restricts a direct neurochemical interpretation, the present microstate findings converge with avalanche and aperiodic results in suggesting more constrained, inhibition-weighted electrophysiological dynamics in our SSD sample.

Lastly, entropy and oscillatory activity (particularly in the beta and gamma range) are other measures suggested to relate to excitation-inhibition^12^. Entropy represents the complexity of signals by measuring randomness in patterns and has been shown to be disrupted in schizophrenia as elevated irregularity or noise in EEG^89,90^. Similarly, we observed an increased beta-range entropy in patients suggesting more unpredictable or disorganized activity, yet a fronto-central lower entropy in the low-frequency range. This differential pattern may reflect a dysregulated spatiotemporal neural hierarchy in SSD. The absence of an association between neurochemical measures and entropy in our study may reflect the fact that entropy is maximized at near-critical states^91^, rather than reflecting a bidirectional measure of excitation-inhibition balance. Similarly, oscillatory activity also showed a mixed pattern. Gamma power attenuation has previously been reported across the schizophrenia illness course, from early-stage psychosis to chronic illness, and has been linked to worse cognition and greater symptom severity^77^. In the present study, we found a reduction of occipital gamma power in the SSD group, although no associations were found with clinical symptoms and cognitive performance. However, recent meta-analytic evidence has shown a reversed pattern of small increase of gamma power in schizophrenia, especially in chronic and medicated patients, although the results across studies are highly variable and dependent on methodological and clinical context^92^. Taken together, in contrast to avalanche criticality and aperiodic activity, oscillatory activity did not provide strong evidence for E-I shifts in the present sample, but may instead reflect regionally specific alterations in neural synchronization that are less directly coupled to frontal MRS metabolite levels investigated in this study^3,12,36^ .

### 4.4 Limitations

Several limitations of the present study should be considered. First, MRS and EEG were not acquired simultaneously. Even though both modalities were collected within a relatively narrow time window (median interval: 0 days), this limits the possibility to investigate real-time influences between neurochemical and electrophysiological dynamics of E-I balance. In addition, potential changes in metabolite concentrations or EEG markers between the two sessions cannot be ruled out, although most recordings were conducted on the same day. Second, the interpretation of some EEG-based E-I measures, particularly oscillatory measures, might have benefited from a high-resolution source-level analysis. However, such analyses were not conducted because the present 32-channel EEG system provides limited spatial resolution and is less suitable for robust source localization than higher-density (>64 channels) EEG setups. Third, the use of a 7T scanner, given its higher signal-to-noise ratio and improved spectral and spatial resolution, would have improved metabolite detection, particularly for resolving metabolites with overlapping peaks, such as glutamate. Lastly, SSD was treated as a single pooled group, which may have obscured disorder-specific mechanisms and relevant heterogeneity within the schizophrenia spectrum. As a result, distinct subgroup patterns or alternative clustering structures may have remained undetected.

## 5. Conclusion

The present study shows widespread alterations in EEG-derived E-I proxies in medicated patients with SSD, pointing out largely suggesting more subcritical, constrained, and inhibition-weighted electrophysiological dynamics. Indication of such inhibition, specifically via avalanche criticality measures, was linked with frontal GABA and Glx/GABA ratio metabolites, as well as with the offset of cognitive and clinical symptoms. Although E-I imbalance is a complex construct that is best understood within a multidimensional framework considering different levels of analysis (e.g., synaptic, local neuronal population, and global network levels), avalanche criticality measures might represent a particularly promising systems-level marker that can capture E-I-related shifts in SSD, with potential relevance for linking neurophysiological alterations to neurochemical mechanisms and clinical outcome.

## Acknowledgements

The MRS package was developed by Edward J. Auerbach and Małgorzata Marjańska and provided by the University of Minnesota under a C2P agreement.

## Declaration of generative AI and AI-assisted technologies in the writing process

During the preparation of this work the authors used the GPT – 5 model developed by OpenAI to improve readability and language of the manuscript. All the authors reviewed and edited the content as needed and take full responsibility for the content of the published article.

## Data and Code Availability

Data and code used for analysis in this paper will be shared by the lead contact upon request.

## Author Contributions

D.K., E.W., and F.J.R. designed and conceptualized the Clinical Deep Phenotyping study. G.H., M.S.K., B.K., V.M., and D.K. formulated the idea and designed the current study. V.M., M.S.K., N.K., M.K., A.H., J.M., and S.S. recruited patients and collected study data. E.W., V.Y., and J.M. trained staff on diagnostic and clinical assessments. MRI measurements were performed by M.S.K., J.M., and S.S. under the supervision of D.K. and L.R. EEG measurements were performed by V.M., N.K., and M.K. under the supervision of D.K. B.K., M.S.K., and G.V. processed MRS data. G.H and B.K. processed EEG data. Statistical analysis and visualization were done by G.H., and B.K. G.H., M.S.K., B.K., and D.K. wrote the manuscript. E.B., M.K., E.S., D.Y., G.V., V.Y., and M.Z., provided critical review. G.H, B.K., M.S.K., and D.K. prepared the final manuscript with the help of all authors. All members of the CDP Working Group contributed to the collection of study data and reviewed the manuscript prior to submission.

## CDP Working Group

Stephanie Behrens, Emanuel Boudriot, Man-Hsin Chang, Valéria de Almeida, Sylvia de Jonge, Fanny Dengl, Peter Falkai, Laura E. Fischer, Nadja Gabellini, Vanessa Gabriel, Sabrina Galinski, Thomas Geyer, Katharina Hanken, Alkomiet Hasan, Genc Hasanaj, Alexandra Hisch, Georgios Ioannou, Iris Jäger, Marcel S. Kallweit, Temmuz Karali, Susanne Karch, Berkhan Karslı, Daniel Keeser, Christoph Kern, Nicole L. Klimas, Maxim Korman, Nikolaos Koutsouleris, Lenka Krcmar, Verena Meisinger, Julian Melcher, Matin Mortazavi, Joanna Moussiopoulou, Karin Neumeier, Frank Padberg, Boris Papazov, Irina Papazova, Sergi Papiol, Pauline Pingen, Oliver Pogarell, Siegfried G. Priglinger, Florian J. Raabe, Lukas Roell, Moritz J. Rossner, Philipp Sämann, Andrea Schmitt, Susanne Schmölz, Eva C. Schulte, Enrico Schulz, Benedikt Schworm, Elias Wagner, Sven Wichert, Vladislav Yakimov, Peter Zill, Zhuanghua Shi, Michael J. Ziller

