## Supplement 1 for "Excitation-Inhibition Balance in Schizophrenia Spectrum Disorders: EEG Criticality Reflects Frontal Metabolites and a Potential Compensatory Mechanism": Supplement1.pdf

Hasanaj, Kallweit *et al.*

### Supplemental Figures

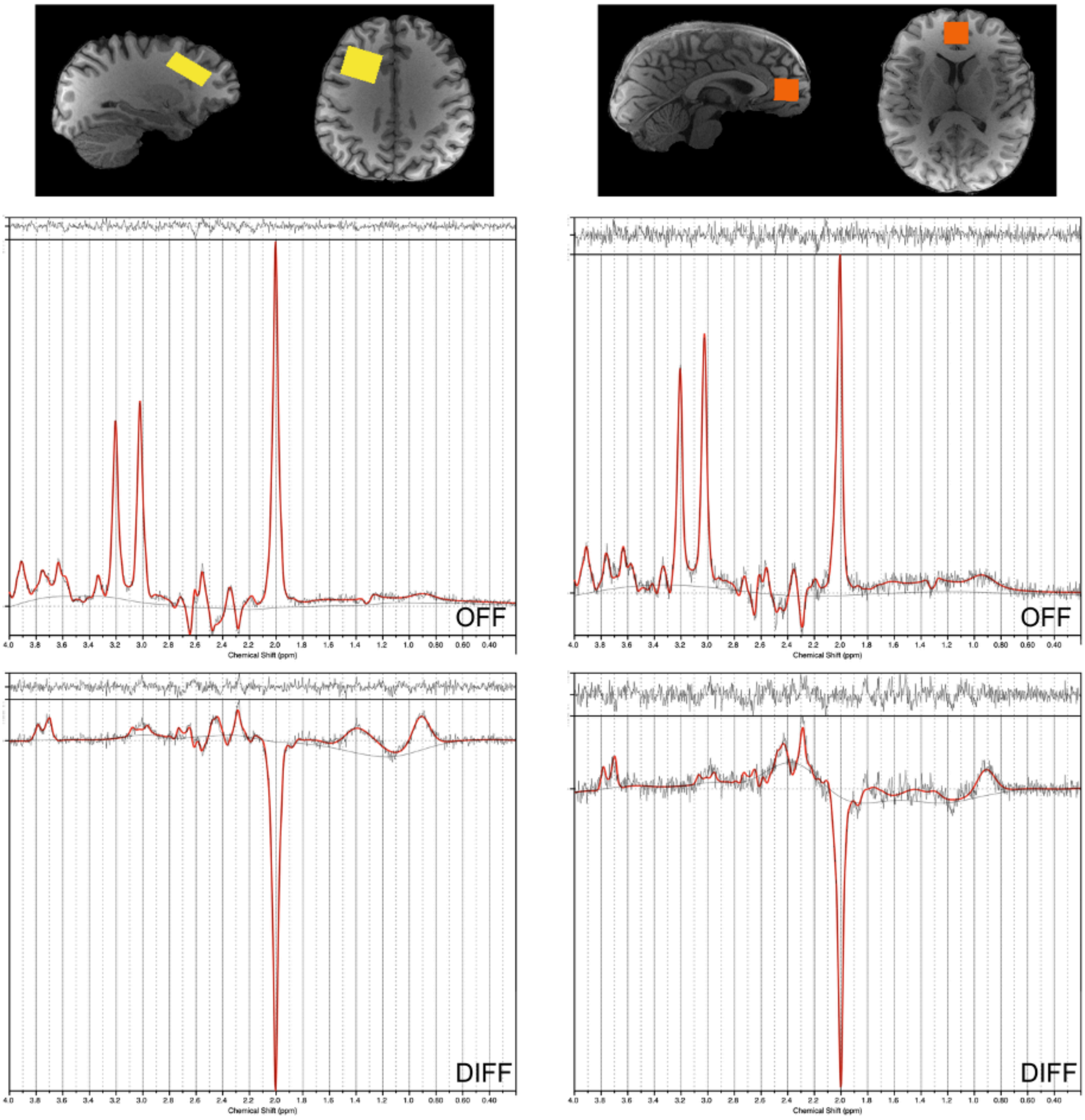


**Figure S1. Typical voxel positioning and associated spectra for the lDLPFC (left) and ACC (right).**
Anatomical sites for MRS voxels are displayed on T1-weighted structural images, complemented by representative unedited (OFF) spectra for Glx quantification and edited (DIFF) spectra for GABA, including their respective LCModel fits.

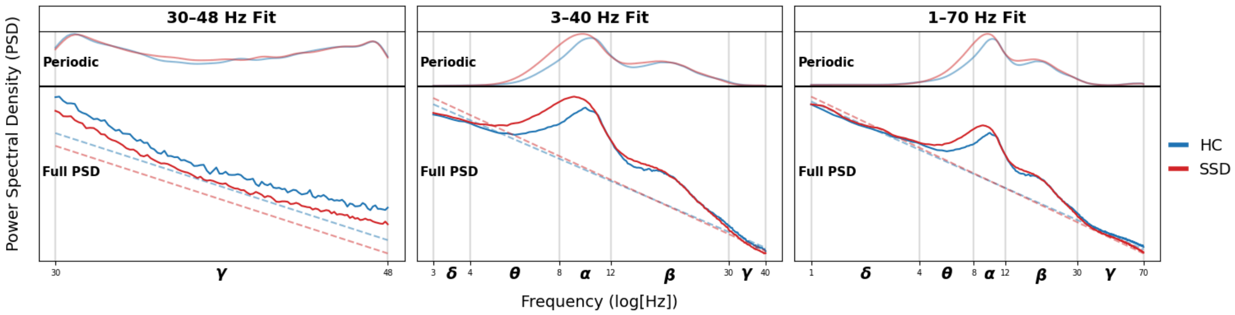


**Figure S2. Group-averaged power spectral density (PSD) decomposition across different fitting ranges.**
The full PSD (solid lines) is shown together with its aperiodic component (dashed lines) and periodic component (top panels). Results are displayed for three fitting ranges: 30–48 Hz, 3–40 Hz, and 1–70 Hz. Blue lines represent healthy controls (HC) and red lines represent individuals with schizophrenia spectrum disorders (SSD). Frequency bands are indicated along the x-axis (δ: Delta, 1–4 Hz; θ: Theta, 4–8 Hz; α: Alpha, 8–12 Hz; β: Beta, 12–30 Hz; γ: Gamma, 30–70 Hz).

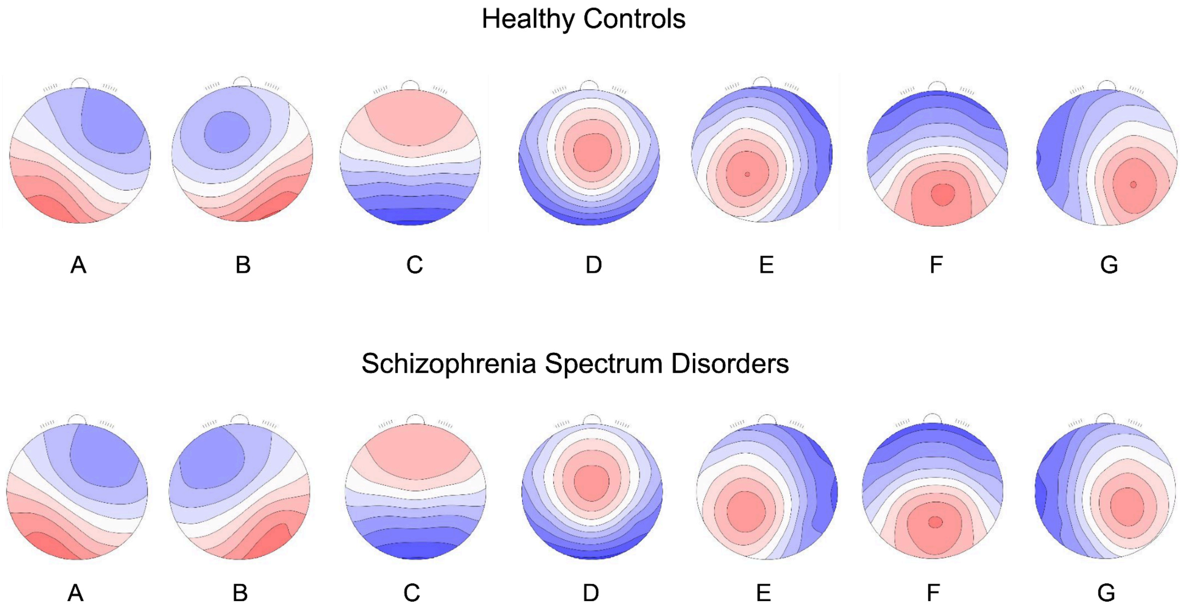


**Figure S3. Group-specific grand mean microstate templates used for backfitting in healthy controls and schizophrenia spectrum disorders.**

Maps A–G show the grand mean scalp topographies obtained separately for each group. These templates were then used for backfitting to individual participants within the corresponding sample.

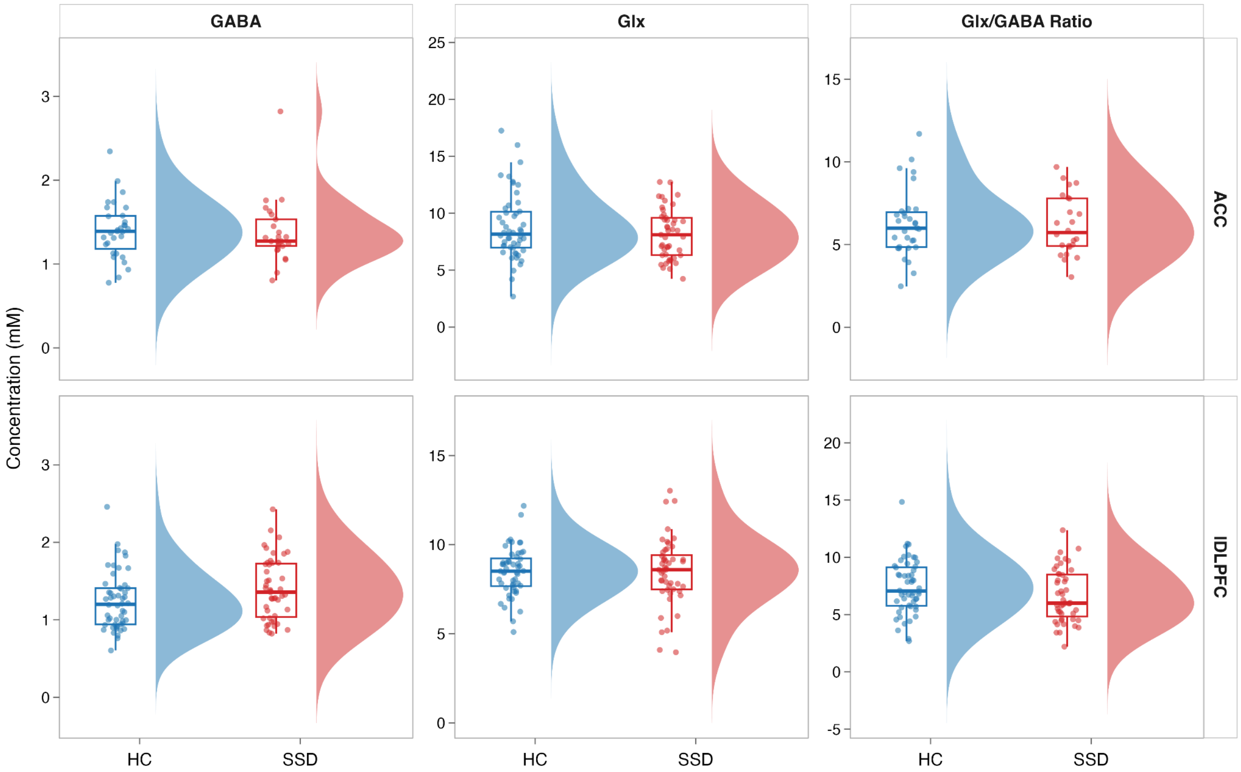


**Figure S4. Regional neurometabolite profiles in healthy controls (HC) and individuals with schizophrenia spectrum disorders.**

The figure shows the distributions of absolute GABA, Glx, and the Glx/GABA ratio in the anterior cingulate cortex (ACC; top row) and left dorsolateral prefrontal cortex (lDLPFC; bottom row) for healthy controls (HC, blue) and participants with schizophrenia spectrum disorder (SSD, red). Each panel includes individual data points, boxplots indicating the median and interquartile range, and half-violin plots illustrating the overall data distribution. No significant group differences were observed for any metabolite measure in either brain region (all $q > 0.05$).

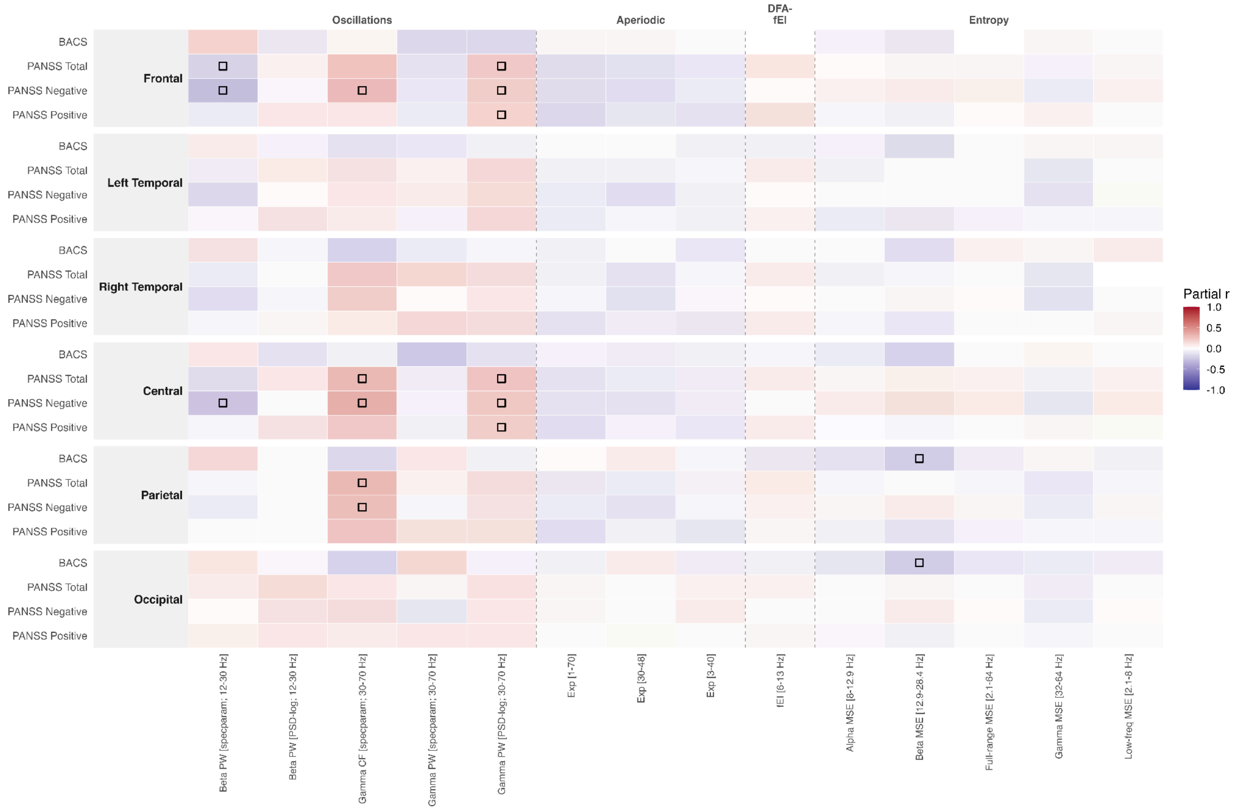


**Figure S5. Electrode-cluster-specific partial correlations of electrophysiological E-I proxies with clinical and cognitive measures in schizophrenia spectrum disorders.**

Rows represent PANSS total, PANSS positive, PANSS negative, and BACS composite z-score within each electrode cluster. Columns include EEG-based measures related to oscillatory activity, aperiodic components, fEI, and multiscale entropy. All partial correlations were adjusted for age, sex, and chlorpromazine-equivalent dose (CPZeq). Color indicates the strength and direction of the partial correlation, with positive values shown in red and negative values shown in blue. Open squares mark nominally significant associations ($p < .05$), and filled squares indicate associations that remained significant after false discovery rate correction ($q < .05$).

### Supplemental Tables

Tables S1-S12: see separate Excel File

### Supplemental Methods

##### EEG data preprocessing

To preprocess the EEG data, we utilized an automated ICA preprocessing pipeline as previously used by Adamd et al.[1]. The whole pipeline is written in MATLAB (The Mathworks Inc.) using EEGLAB[2] v2025.0; https://sccn.ucsd.edu/eeglab/). EEG data were re-referenced to the mastoid electrodes A1 and A2 and downsampled to 256 Hz. Data were then band-pass filtered between 1 and 70 Hz, with an additional notch filter applied between 49.5 and 50.5 Hz to remove line noise. Continuous data were subsequently segmented into 6-second epochs.

Artifact rejection followed a multi-step procedure. Epochs with a mean amplitude exceeding ±5 standard deviations were removed. Additional epoch rejection was performed to exclude segments containing linear trends using pop_rejtrend with a maximum slope of 5 and a minimum $R^2$ of 0.7. Spectral rejection was applied using pop_rejspec, excluding epochs with low-frequency activity below 2 Hz exceeding a power threshold of -50 to 50 dB, and high-frequency activity between 20 and 40 Hz exceeding a power threshold of -100 to 25 dB. Bad channels were identified using pop_rejchan based on extreme values in the power spectrum, kurtosis, and joint probability. The applied thresholds were -4 to 6 standard deviations for the power spectrum, -7 to 15 standard deviations for kurtosis, and -9 to 7 standard deviations for joint probability. If more than 50% of epochs were rejected at this stage, the rejection sequence was repeated for the initial data with channel rejection performed before epoch rejection. Datasets were excluded from further analysis if, after this repeated procedure, more than 50% of the data remained rejected or more than 20% of electrodes were removed.

For datasets passing these quality-control criteria, preprocessing continued with Independent Component Analysis (ICA). Artifact-related independent components were identified and removed using the Multiple Artifact Rejection Algorithm[3] (MARA). Following MARA correction, a second round of epoch and channel rejection was applied, with slightly adjusted channel rejection thresholds of -6 to 5 standard deviations for the power spectrum, −6 to 9 standard deviations for kurtosis, and -7 to 7 standard deviations for joint probability. Bad or missing channels were then interpolated, and the epoched data were converted back to continuous data using eeg_eeg2continuous for subsequent analyses. The preprocessing of the EEG data was conducted on the whole parent CDP project[4], and included in the present study are only subjects that had both EEG and MRS data.

**EEG E-I Metric Calculation**

In the present study, we extracted seven different EEG markers to track E-I brain dynamics, as were originally proposed by Ahmad et al. [5]

I. Gamma Power and Center Frequency: The Power Spectrum Density (PSD) was estimated using the Welch method (6-second windows, 50% overlap) implemented in MNE-Python[6] (v1.8.0), which was then further preprocessed using the *specparam* algorithm to isolate the oscillatory (periodic) activity from the aperiodic activity[7] (https://github.com/fooof-tools/fooof). Both gamma power and center frequency were extracted after applying the *specparam* algorithm. The model used to extract gamma power and center frequency was a fitting range of 1–70 Hz, peak width limit: [1, 12], minimum number of peaks: 8, peak threshold: 1, and minimum peak height: 0.1, using the "fixed" fitting method (Figure S1 in Supplement 1). The gamma power was then extracted for power and center frequency of peaks within 30–70 Hz. Given that many previous studies have not accounted for 1/f aperiodic contributions, we also included canonical estimates of absolute gamma power for comparison.

II. Beta Power: Was extracted from the same *specparam* fitted model of 1–70 Hz, from peaks within the 12–30 Hz range. As with gamma, we also included canonical absolute beta power in our analyses to account for potential differences in methodological approaches across studies.

III. Aperiodic Exponent: We calculated three aperiodic exponents that are commonly used in the literature. The first was derived from a *specparam* model fit across a 3–40 Hz frequency range to replicate our previous analysis in the same CDP cohort[1]. The second exponent was fitted in the lower gamma range [30–48 Hz], to approximate the originally proposed range (30–50 Hz) recommended by Gao et al.[8]. The last exponent was fitted in the broader 1–70 Hz, from which we also extracted the aperiodic-adjusted oscillatory activity for both beta and gamma power. We used the "fixed" fitting approach for all the models (Figure S2 in Supplement 1).

IV. Functional E-I (fEI): fEI was computed using a detrended fluctuation analysis (DFA) method, originally introduced by Bruining et al.[9] (2020), applied to the extended alpha frequency range (6–13 Hz). Following the previous applied criteria[10], we included only fEI values associated with DFA values between 0.55 and 1.

V. Microstate Dynamics: To address variability across studies and improve explained variance, we used the seven-class solution proposed by Custo et al.,[11] that can better separate correlated topographies with identifiable distinct sources (e.g., microstate C and F). EEG data were bandpass-filtered from 2–20 Hz, consistent with standard microstate preprocessing. The MICROSTATELAB[12] plugin in EEGLAB was used to calculate the microstate dynamics. For each of the seven classes, we computed the mean duration and mean occurrence, leading to a total of 14 variables. For each group (HC and SSD), we backfitted the corresponding group-specific grand mean (Figure S3 in Supplement 1). The topography of each microstate was visually inspected for each participant, and included in further analyses if it matched the typical microstate topography.

VI. Entropy: Multiscale sample entropy (MSE) was calculated for the whole EEG signal at each electrode, using the nk.entropy_multiscale function from the NeuroKit2[13] Python library (https://neuropsychology.github.io/NeuroKit/). MSE is a nonlinear method that estimates entropy by coarse-graining the original time series at increasing scale factors, thereby progressively smoothing the data. This approach enables frequency-sensitive analysis, as smaller scales retain faster (high-frequency) dynamics, whereas larger scales increasingly reflect slower (low-frequency) components of the signal[14]. The MSE was calculated by setting an embedding dimension (m) to 2, and the tolerance (r) at 0.2, to keep consistency with previous studies in SSD[15,16]. We extracted entropy measures from five scale ranges, each approximating the respective frequency band: full-range (scales 4–120; ~64–2.1 Hz), low-frequency (32–120; ~8–2.1 Hz), alpha (21–32; ~12.9–8 Hz), beta (9–21; ~28.4–12.9 Hz), and gamma (4–8; ~64–32 Hz).

VII. Neuronal avalanches: To calculate neuronal avalanches, we applied z-score normalization to the EEG data and used log-density distribution plots to determine an event-detection threshold, noting deviations from the Gaussian distribution at approximately ±2.5 SD. Neuronal avalanches were then extracted following Lombardi[17] et al., and we identified a bin size of 7 as optimal, as it yielded a power-law distribution of avalanche sizes with an exponent ($\alpha$) closest to -3/2 in healthy controls, indicative of criticality[18]. We further calculated the branching ratio and avalanche exponent to assess network excitability, and computed the kappa index to evaluate the goodness of fit to the theoretical $\alpha = -3/2$ distribution based on Shew et al.[19]
